# Cost-effectiveness analysis of two-way texting (2wT) intervention to improve ART retention among newly-initiated antiretroviral therapy clients in Malawi

**DOI:** 10.1101/2024.04.17.24305960

**Authors:** Christine Kiruthu-Kamamia, Hiwot Weldemariam, Mirriam Chipanda, Jacqueline Huwa, Johnnie Seyani, Harrison Chirwa, Aubrey Kudzala, Agnes Thawani, Joseph Chintedza, Odala Sande, Geldert Chiwaya, Hannock Tweya, Milena Pavlova, Wim Groot, Caryl Feldacker

## Abstract

**Background:** Retention in HIV care is crucial for improved health outcomes. Malawi has a high HIV prevalence and struggles with retention despite significant progress in controlling the epidemic. Mobile health (mHealth) interventions, such as two-way texting (2wT), have shown promise in improving anti-retroviral therapy (ART) retention. We explore the cost-effectiveness of a 2wT intervention in Lighthouse Trust’s Martin Preuss Center (MPC) in Lilongwe, Malawi, that sends automated SMS visit reminders, weekly motivational messages, and supports direct communication between clients and healthcare workers.

**Methods:** Costs and retention rates were compared between 2wT and standard of care (SOC) for 468 clients enrolled in each. Incremental cost-effectiveness ratios (ICERs) were calculated. Scenario analyses were conducted to estimate costs if 2wT expanded.

**Results:** The 2wT group had higher retention (80%) than SOC (67%) at 12 months post-ART initiation. For 468 clients, the total annual costs for 2wT were $36,670.38 as compared to SOC costs at $33,458.72, resulting in an ICER of $24,705. Among scenarios, the ICER was -$105,315 if 2wT expanded to all new clients (2678 at MPC and -$723,739 as 2wT expanded to other four high-burden facilities (2901 clients), suggesting high cost savings if 2wT was effectively scaled.

**Conclusion:** The 2wT intervention appears cost-effective to improve ART retention among new ART initiates in a high-burden ART clinic. While mHealth interventions have potential limitations, their benefits in improving patient outcomes and cost savings support their integration into HIV care programs.

## Background

Retention on antiretroviral therapy (ART) among people living with HIV (PLHIV) leads to lower mortality and a higher likelihood of viral load (VL) suppression, thereby reducing the risk of HIV transmission [1,2]. However, ART retention continues to be a major challenge [3,4], especially within the first 12 months of ART initiation as research, including studies conducted in Malawi has shown that the chances of attrition are highest within the first year of initiating ART [5–9]. Malawi has one of the highest HIV prevalence in the world, with approximately 9% of the general population living with HIV [10]. Malawi adopted a public health approach with the aim to achieve UNAIDS 95-95-95 targets by 2030 [11]. Progress is commendable: by 2021, 88% of those living with HIV knew their status, 98% of those were on ART, and 97% had their VL suppressed [10]. Despite this significant achievement, Malawi continues to struggle with retention and adherence to ART [12,13].

Generally, adherence is influenced by several factors, including predisposing factors (e.g., mental illness, substance abuse), patient enabling factors (e.g., reminder strategies, transportation) and healthcare environment factors (e.g., provider characteristics, clinic experience [14]. To address these challenges, the World Health Organization (WHO) recommended evidence-based interventions, including mobile health (mHealth) approaches like reminders [15].

Increased access to mobile phones and their rapid technological advancements have led to the development of mHealth as a complementary strategy to strengthen health systems [15]. mHealth is a medical and public health practice that is supported by mobile devices, patient monitoring devices, personal digital assistants, and other wireless devices [15]. mHealth is a growing strategy with over 600 pilots and programs implemented globally over the last decade [16]. Specific mHealth innovations have shown promise in increasing ART retention in research settings [17].

The use of text messaging, like short messaging service (SMS), for patients and their health providers to communicate has been steadily increasing due convenience, accessibility, and privacy advantages [17,18]. Studies on SMS communication have found that interactive two-way SMS (sending messages with response options) between patients and health providers is more efficacious than one-way informational messaging (sending messages that do not require a response) because it facilitates interaction between the provider and the client [19–21]. SMS can be used to promote adherence by sending prompts to take HIV medication, appointment reminders, and interacting with healthcare providers, with results from Kenya, Burkina Faso and Nigeria showing improved uptake and adherence[17,19,22,23]. Furthermore, a meta-analysis by Wald et al found that two-way messaging was associated with substantially improved medication adherence, compared to one-way text messaging [20]. Based on such evidence, the WHO recommended SMS messaging as an intervention to promote ART [24]. Previous research suggests that two-way SMS interventions could be cost-effective [25,26], but evidence gaps remain. As mHealth interventions continue to grow exponentially, there is a critical gap in evaluation and evidence generation to scale only cost-effective, impactful innovations.

Lighthouse Trust (LH) is a WHO-recognized center of excellence for HIV care that has been operational since 2001 [27–29]. LH operates 5 clinics in Malawi: two in the central district Lilongwe (Lighthouse Trust at Kamuzu Central Hospital, Martin Preuss Center at Bwaila Hospital), two in the southern districts Blantyre (Umodzi Family Center at Queen Elizabeth Central Hospital) and Zomba (Tisungane Clinic at Zomba Central Hospital) and one in the northern district Mzimba (Rainbow Clinic at Mzuzu Central Hospital) [30].

Lighthouse Trust’s Martin Preuss Center (MPC) clinic in Lilongwe, Malawi has the largest ART cohort in the country, with over 25,000 clients alive in care [31]. At MPC, there are over 7,800 monthly visits to the clinic. Approximately 10% of clients at MPC miss their appointments each month. MPC has a policy of following up on those who miss their appointment, which means, on average, 780 clients need tracing monthly. Tracing is done by field tracers who trace clients telephonically or physically to their homes. With such a high demand for client tracing, coupled with limited resources, only about one-third of eligible clients are traced. Lack of, or delays in, tracing reduces ART retention and, ultimately, viral suppression. Poor data quality also hampers tracing effectiveness. On average, of all clients traced, approximately 55% were not loss-to-follow-up (LTFU) but had transferred to another clinic or were actually still in care, resulting in the inefficient allocation of limited resources [13].

To address this problem, Lighthouse Trust collaborated with the International Training and Education Center for Health (I-TECH) at the University of Washington, along with technology partner, Medic, leveraged the open-source Community Health Toolkit (CHT) [32], to customize the two-way texting (2wT) system to enhance early retention among new ART initiates. The 2wT system sends automated visit reminders and weekly motivational messages to clients. Clients can respond to inquiries and send messages to the HCW to change visit dates, report a transfer, or ask for visit-related help. 2wT is free for clients. By directly, and proactively, engaging with clients before a visit is missed, 2wT aimed to improve client outcomes, reducing true LTFU. By identifying transfers and delays before a visit was missed, 2wT aimed to reduce wasted tracing efforts. Six-month retention analysis demonstrated that the 2wT system improved ART retention by 10% within the first six months of ART, while also lowering the risk of LTFU by 64% compared to the standard of care (SOC) approach (forthcoming). Despite initial effectiveness evidence, the cost and cost-effectiveness of this mHealth intervention remain unknown.

Therefore, the primary objective of this study is to assess the cost of implementing the 2wT mHealth intervention and the cost-effectiveness of the intervention in comparison with the standard of care (SOC) buddy approach (visit reminder calls from an expert ART *buddy)* at MPC ART clinic in Lilongwe. In response to recent calls for evidence on SMS innovations and cost-effective interventions from the Malawi Ministry of Health (MOH) [12], this cost and cost-effectiveness analysis from the program perspective may inform feasibility of 2wT scale-up to other MoH facilities.

## Methods

### Study design

This study was based in the Lighthouse Trust MPC ART clinic in Lilongwe, Malawi. Using a program (payer) perspective, we conducted a cost-effectiveness analysis comparing 2wT intervention to the SOC. Specifically, we evaluated the costs incurred by the clinic implementing and providing routine retention services (SOC) and the effect of the intervention (2wT) in improving client retention. The base case (SOC) included adult clients with mobile phones who initiated ART at MPC between January and December 2020. SOC clients were supported with *Buddy* reminder calls before a visit and after a missed appointment, if applicable. 2wT clients were also adult clients with cell phones who opted into enrollment in the 2wT intervention from June 2021 to April 2022. We calculated the costs of early retention in 2020-2021. All client outcomes were followed up to 12 months post-ART initiation. The early retention procedure is presented in Figure 1. The economic evaluation was conducted following the Consolidated Health Economic Evaluation Reporting Standards 2022 (CHEERS 2022)[33], Appendix 2.

**Figure 1:**
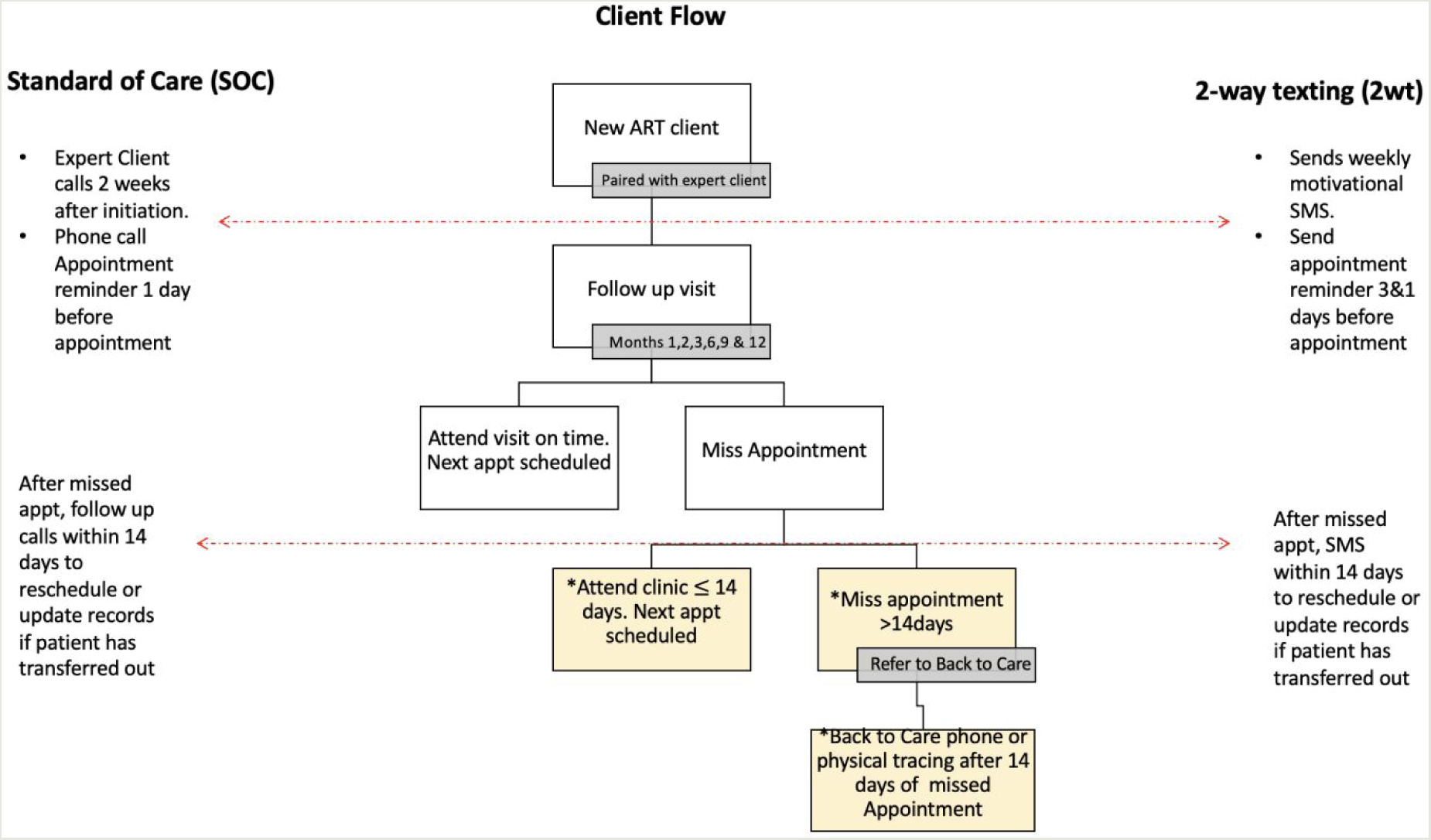
Early retention procedures for new ART clients in SOC and 2wT. Note: After a client misses an appointment, the procedures are the same for all clients regardless of the intervention and when they initiated care.

### Standard of care (SOC)

For client retention, Lighthouse implements a resource-intensive patient retention program that is split between early retention for new ART initiates (within the first 12 months of ART initiation) and retention of long-term clients (those on ART longer than 1 year) called Back to Care (B2C). In the early retention program (see Figure 1), a newly initiated ART client must have ART clinic visits at months 1, 2, 3, 6, 9 and 12 post-initiation for close observation. In addition, new ART clients are paired with an expert client buddy (EC), who are peer counsellors living with HIV. This pairing is done for the first 12 months of ART initiation. The EC, in addition to providing peer counselling, calls the client for appointment reminders before their scheduled appointment and traces clients through phone calls within 14 days after a missed appointment. If phone tracing is unsuccessful within 14 days and the client has not attended the clinic, the case is sent to B2C for additional tracing, which includes phone calls or home visits. The early retention register documents all phone call reminders and tracing attempts. However, human resource challenges and documentation gaps resulted in only 6 months of recorded buddy data per client.

### The two-way texting (2wT) intervention

Previously described elsewhere [34], the 2wT system replaced phone calls made by ECs for counseling, appointment reminders and tracing. Clients who fulfilled the eligibility requirements were offered the opportunity to provide their consent and opt-in the 2wT study if they: 1) had initiated ART within the past six months; 2) were aged 18 years or older; 3) possessed a phone at the time of enrollment; 4) expressed willingness to receive and send SMS messages; 5) had literacy skills; and 6) acknowledged and verified their enrollment phone number by receiving and confirming the 2WT enrollment SMS. Clients who either did not have cell phones or had but chose not to participate were excluded from the study and instead received SOC retention support. The 2wT uses a hybrid workflow that combines: (1) automated workflows that send weekly motivational messages to promote adherence; (2) individually-tailored SMS reminders to clients with upcoming visits, with a response requested; and (3) open-ended SMS texts between clients and HCWs that allows clients to reschedule visits, report transfers, or report other clinical or non-clinical interactions such as ART side effects or ask questions on their ART care. The system is used for the first 12 months of ART initiation. Appointment reminders are sent 3 and 1 days before the appointment by automated SMS. After a missed appointment, automated SMS reminders are sent within 14 days to reschedule the appointment or update records if the client has transferred to another clinic. If a client enrolled in 2wT has not attended the clinic within 14 days after a missed appointment, the case is sent to B2C for in-person tracing as with SOC. Participants in 2wT did not receive an EC buddy. For both 2wT and SOC, if a client has not returned to the clinic within 14 days after a missed appointment, the case is referred to B2C for additional tracing

We hypothesize that 2wT is more cost-effective for a high-burden facility such as MPC.

### Ethics

For the underlying 2wT implementation effectiveness study, 2wT patients provided written informed consent at opt-in enrollment in either Chichewa or English according to participant preference. The study protocol was approved by the Malawi National Health Sciences Research Committee and the University of Washington, Seattle, USA ethics review board. No identifiable data was used in this costing study.

### Data Collection

We conducted a time-motion study to record staff time spent on SOC retention and 2wT activities. The data collection tools were developed using Microsoft Excel, provided in Appendix 1. The time and motion study consisted of five days of direct observation at MPC.

#### Outcomes (effects)

The main clinical outcome evaluated was reduced clients defaulted at 12 months post ART initiation. Client ART outcome data from both 2wT and comparison clients were extracted from the EMR, EC tracing registers, and the 2wT database. ART outcome data came from the EMRs, and the SMS data from the 2wT database. The tracing attempts by ECs were documented in the early retention register.

#### Costs

We used a micro-costing approach for cost estimation. Costing information was obtained from Lighthouse Trust expenditure records, payroll information and procurement records, and the time motion surveys designed for the study. We identified and categorized all activities and resources involved in both the SOC and 2wT intervention for early retention. These costs were categorized into personnel, training, building utilities, supplies, equipment, and communication materials as described in Table 1. The costs were further divided into fixed costs and variable costs. The fixed costs were one-time expenses, which included training and equipment costs. The fixed costs were allocated to SOC and 2wT both interventions based on their proportional utilization of shared resources. The variable costs included recurring costs required to sustain both interventions. These included personnel, communication materials, supplies and building utilities.

**Table 1:** Cost Categories.

| Cost Category | Description |
| --- | --- |
| **Personnel** | Value of personnel time and effort spent in each activity |
| **Equipment** | Investments that last longer than 1 year, including mobile phones, laptops, and furniture |
| **General supplies** | Supplies used for documentation and contacting clients |
| **Training** | Expenses used in training sessions for 2wT use and SOC |
| **Communication** | Costs for phone service companies |
| **Utilities** | Costs for utilities including electricity, water, and internet |

Note: SOC, standard of care; 2wT, two-way texting

We quantified and valued the resources for each cost category using Lighthouse expenditure data on salaries and commodity prices. Since the perspective of the analysis was Lighthouse Trust (payer) perspective, we excluded costs that are not incurred by the clinic, such as medication costs, which are paid by the government, and study-specific personnel that would not be transferable to routine program implementation.

### Discounting

We used 5% social discount rate as recommended for LMICs due to higher rates of economic growth [35].

### Currency, price date, and conversion

All costs were converted from Malawi Kwacha (MWK) to US dollar. We used the 2020-2021 exchange rate of 825 MWK to match the study period of the client outcomes.

### Assumptions

*We had the following assumptions*.

1. We assumed the program all staff were working 40 hours/week.
2. The annual costs for SOC are for all 2,678 clients seen in 2020 and could not be easily extracted for 468 clients, only. Therefore, to estimate costs for the 468 clients in SOC, we estimated all personnel and communication costs needed for each client, multiplying that by 468.

### Data analysis

We analyzed the 12 -month ART outcomes for each client and estimated the costs per client enrolled in SOC and 2wT and retained at 12-months. We calculated the incremental cost effectiveness ratios (ICERs) of 2wT compared to the SOC [36].

### Sensitivity and Scenario Analyses

Sensitivity analyses were conducted to investigate how the ICER of the SOC and 2wT changed if 2wT was scaled beyond the pilot, making it accessible to all new ART clients at MPC, and if it was expanded beyond MPC to other facilities. We calculated the ICERs for both scenarios.

Scenario 1 (scale up to all new ART initiates at MPC): We assumed that 44% of all new initiates at MPC in 2020 would be eligible for 2wT (according to enrollment screening data). Therefore, of the 2,678 initiated on ART at MPC in 2021, we estimated the cost of providing retention support to 1,005 in SOC and 942 clients in 2wT. In the SOC, the personnel and communication costs were estimated by multiplying the unit cost per client of the base case by the number of clients enrolled. The supplies, utilities, training, and equipment would remain the same. It is estimated that one full-time FTE 2wT data officer can manage up to 3000 patients.

Scenario 2 (scale up to 4 additional facilities): The second scenario estimated costs if 2wT was scaled to the other four LH high-burden ART facilities across the country which enrolled 2,901 new ART clients in 2022. It is estimated that the existing 2wT data officer and a retention assistant (RA) would be enough to manage the 2wT system and client enrolment in this scenario. The enrollment eligibility was presumed the same as from screening data. For personnel, communication, and supplies cost, we multiplied the unit cost per client from the base case by the number of clients). For personnel, communication, and supplies costs, we multiplied the unit cost per client from the base case by the number of clients. We assumed the utilities, training, and equipment costs would be the same for each facility. For 2wT, one data officer from MPC would serve multiple sites, but each facility would use an EC at 20% FTE for enrolling new clients duty, only. For communication and supplies costs, we multiplied the unit cost per client from the base case by the number of clients. Each facility would have the same equipment costs, excluding a lockable cabinet and 2wT system maintenance costs since the 2wT system would be stationed at MPC. For training costs, the ECs would only need 2wT-specific training, but not retention program training, since they would have already received that as part of their normal duties.

## Results

### Demographics and retention outcomes

Table 2 presents the baseline characteristics and 12-month ART outcomes of patients included in SOC and 2wT. Retrospectively, 468 adult new ART clients aged 18 and over with a registered mobile phone number were randomly selected in SOC for comparison with the number of 468 participants who were enrolled in the 2wT intervention. As shown in Table 2, in both SOC and 2wT intervention groups, there were more women than men (55% and 56%, respectively) and the median age was 33 years, with more participants in the 25-34 age group. The majority (77% and 78% at SOC and 2wT respectively) of the participants were initiated on ART at WHO stage 1 or 2, reflecting the demographic profile of new ART clients at MPC[31]. At 12 months post-ART initiation, 2wT intervention had more (80%) clients alive in care compared to those in the SOC (67%). In addition, SOC had more clients loss-to-follow-up (LTFU) (17% vs 6%), and a higher rate of clients who stopped ART (7% vs 2%) compared to the 2wT. The transfer-out rate and death rate were the same for both interventions at 8% and 1% respectively.

**Table 2:**
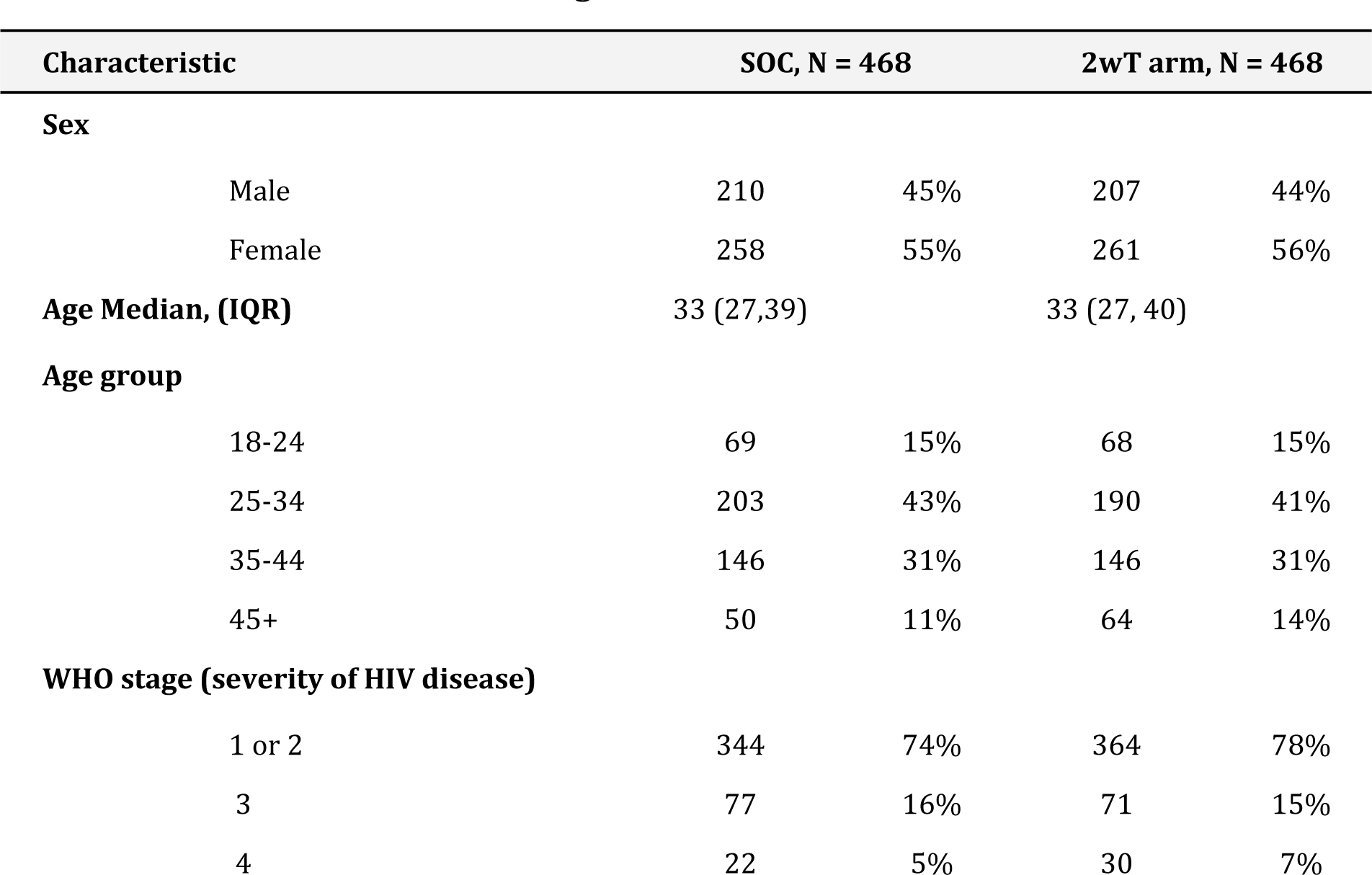
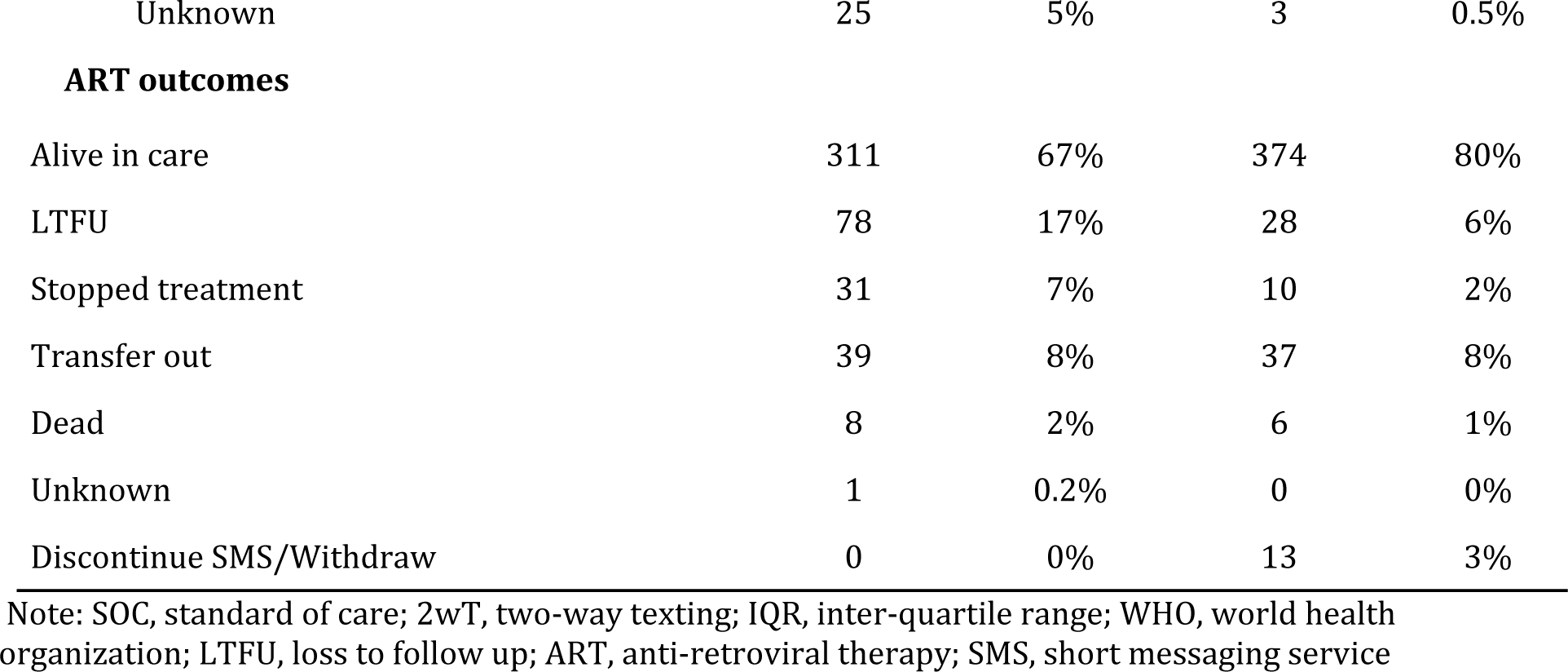
Baseline characteristics and 12-month ART outcomes of patients included in SOC and 2wT at Martin Preuss Centre, Lilongwe, Malawi.

| Characteristic | SOC, N = 468 |  | 2wT arm, N = 468 |  |
| --- | --- | --- | --- | --- |
| **Sex** |  |  |  |  |
| Male | 210 | 45% | 207 | 44% |
| Female | 258 | 55% | 261 | 56% |
| **Age Median, (IQR)** | 33 (27,39) |  | 33 (27, 40) |  |
| **Age group** |  |  |  |  |
| 18-24 | 69 | 15% | 68 | 15% |
| 25-34 | 203 | 43% | 190 | 41% |
| 35-44 | 146 | 31% | 146 | 31% |
| 45+ | 50 | 11% | 64 | 14% |
| **WHO stage (severity of HIV disease)** |  |  |  |  |
| 1 or 2 | 344 | 74% | 364 | 78% |
| 3 | 77 | 16% | 71 | 15% |
| 4 | 22 | 5% | 30 | 7% |
| Unknown | 25 | 5% | 3 | 0.5% |
| ART outcomes |  |  |  |  |
| Alive in care | 311 | 67% | 374 | 80% |
| LTFU | 78 | 17% | 28 | 6% |
| Stopped treatment | 31 | 7% | 10 | 2% |
| Transfer out | 39 | 8% | 37 | 8% |
| Dead | 8 | 2% | 6 | 1% |
| Unknown | 1 | 0.2% | 0 | 0% |
| Discontinue SMS/Withdraw | 0 | 0% | 13 | 3% |

Note: SOC, standard of care; 2wT, two-way texting; IQR, inter-quartile range; WHO, world health organization; LTFU, loss to follow up; ART, anti-retroviral therapy; SMS, short messaging service

### Costs

Table 3 summarizes the total annual costs and unit costs for implementing SOC services and the 2wT intervention. The annual costs for 2wT were higher at $36,670.38 compared to $33,458.72 for SOC. Both fixed costs and variable costs were greater for 2wT. Personnel costs constituted the highest expense for 2wT, while the building costs were the highest expense for SOC. Overall, the cost per client enrolled in the study was higher for 2wT at $78.36 compared to $71.49 for SOC, with a difference of $6.87 between the two. However, the cost per client retained in care after 12 months was higher for SOC ($107.58 vs $98.0) due to more clients retained in care for 2wT.

**Table 3:** Total annual costs and unit costs of SOC and 2wT.

| Categories | Total cost |  | Cost per client |  | Cost per client retained at 12mos |  |
| --- | --- | --- | --- | --- | --- | --- |
|  | SOC | 2WT | SOC, N=468 | 2WT, 468 | SOC, N=311 | 2wt, N=374 |
| Fixed Costs (one-time) |  |  |  |  |  |  |
| Training | \$6,592.18 | \$9,285.32 | \$14.09 | \$19.84 | \$21.20 | \$24.83 |
| Equipment | \$3,603.03 | \$8,909.44 | \$7.70 | \$19.04 | \$11.59 | \$23.82 |
| Variable Costs (recurrent) |  |  |  |  |  |  |
| Personnel | \$9,597.10 | \$15,187.17 | \$20.51 | \$32.45 | \$30.86 | \$40.61 |
| Supplies | \$4,743.15 | \$111.15 | \$10.13 | \$0.24 | \$15.25 | \$0.30 |
| Communication | \$87.27 | \$1,395.37 | \$0.19 | \$2.98 | \$0.28 | \$3.73 |
| Utilities | \$8,835.99 | \$1,781.92 | \$18.88 | \$3.81 | \$28.41 | \$4.76 |
| Total | \$33,458.72 | \$36,670.38 | \$71.49 | \$78.36 | \$107.58 | \$98.05 |

Note: SOC, standard of care; 2wT, two-way texting

We also assessed the costs per gender, age group and WHO stage. Notably, in general, the costs were higher for 2wT in all categories except for the unknown WHO stage.

The cost shares by input categories for all costs are shown in Appendix 3. The personnel, training, equipment, and communication costs were higher for 2wT at 41%, 25%, 24% and 4%, respectively, compared to the respective allocations of 29%, 20%, 11% and 0.3% for SOC. Conversely, the utilities and general supplies were higher for SOC services, with 26%, and 14%, respectively, in contrast to 5% and 0.3% for 2wT.

### Scenario analysis

Tables 4 and 5 present the annual and unit costs for SOC and 2wT for scenarios 1 and 2, respectively.

**Table 4:**
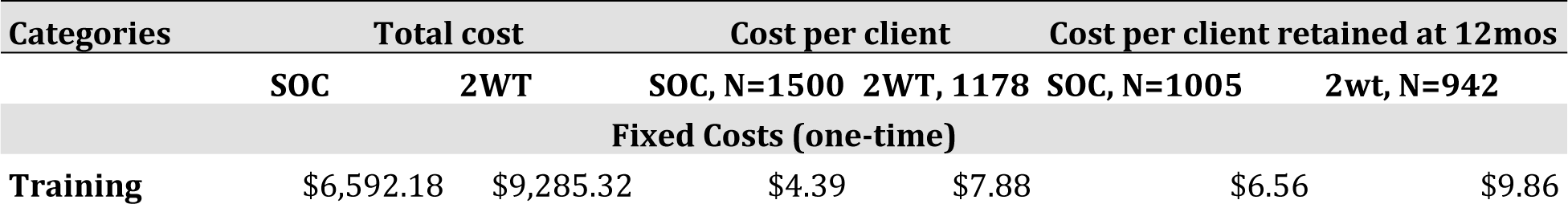
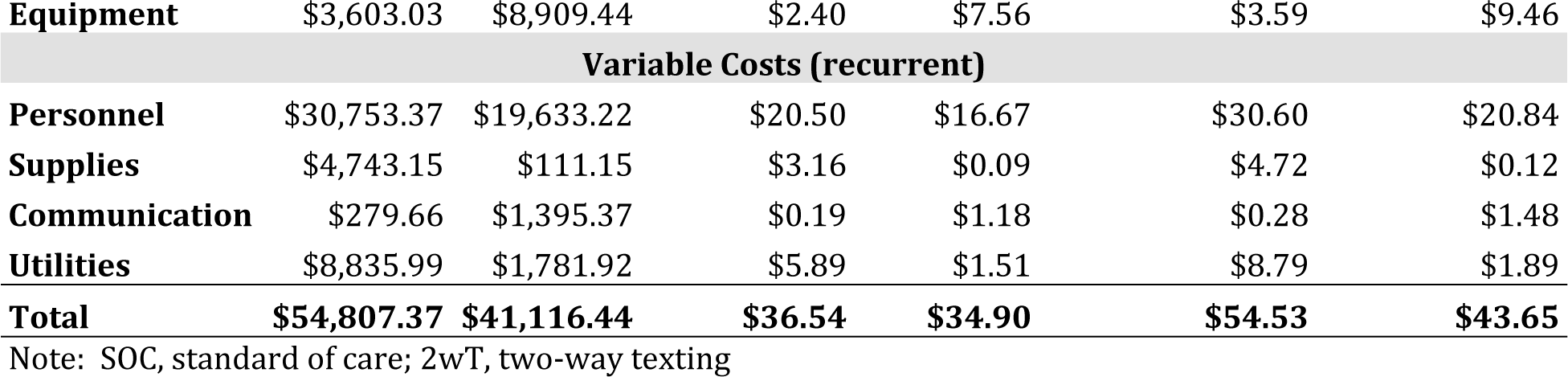
Scenario 1 – total and unit costs of SOC and 2wT.

| Categories | Total cost |  | Cost per client |  | Cost per client retained at 12mos |  |
| --- | --- | --- | --- | --- | --- | --- |
|  | SOC | 2WT | SOC, N=1500 | 2WT, 1178 | SOC, N=1005 | 2wt, N=942 |
| Fixed Costs (one-time) |  |  |  |  |  |  |
| Training | \$6,592.18 | \$9,285.32 | \$4.39 | \$7.88 | \$6.56 | \$9.86 |
| **Equipment** | \$3,603.03 | \$8,909.44 | \$2.40 | \$7.56 | \$3.59 | \$9.46 |
| **Variable Costs (recurrent)** |  |  |  |  |  |  |
| **Personnel** | \$30,753.37 | \$19,633.22 | \$20.50 | \$16.67 | \$30.60 | \$20.84 |
| **Supplies** | \$4,743.15 | \$111.15 | \$3.16 | \$0.09 | \$4.72 | \$0.12 |
| **Communication** | \$279.66 | \$1,395.37 | \$0.19 | \$1.18 | \$0.28 | \$1.48 |
| **Utilities** | \$8,835.99 | \$1,781.92 | \$5.89 | \$1.51 | \$8.79 | \$1.89 |
| **Total** | **\$54,807.37** | **\$41,116.44** | **\$36.54** | **\$34.90** | **\$54.53** | **\$43.65** |

Note: SOC, standard of care; 2wT, two-way texting

**Table 5:** Scenario 2 – total and unit financial costs of SOC and 2wT.

| Categories | Total cost |  | Cost per client |  | Cost per client retained at 12mos |  |
| --- | --- | --- | --- | --- | --- | --- |
|  | SOC | 2WT | SOC, N=1625 | 2WT, 1276 | SOC, N=1121 | 2wt, N=1021 |
| **Fixed Costs (one-time)** |  |  |  |  |  |  |
| **Training** | \$26,368.74 | \$10,772.56 | \$16.23 | \$8.44 | \$23.52 | \$10.55 |
| **Equipment** | \$14,412.10 | \$3,802.60 | \$8.87 | \$2.98 | \$12.86 | \$3.72 |
| **Variable Costs (recurrent)** |  |  |  |  |  |  |
| **Personnel** | \$33,314.23 | \$13,436.61 | \$20.50 | \$10.53 | \$29.72 | \$13.16 |
| **Supplies** | \$16,464.82 | \$303.16 | \$10.13 | \$0.24 | \$14.69 | \$0.30 |
| **Communication** | \$302.95 | \$3,805.79 | \$0.19 | \$2.98 | \$0.27 | \$3.73 |
| **Utilities** | \$35,343.94 | \$0.00 | \$21.75 | \$0.00 | \$31.53 | \$0.00 |
| **Total** | **\$126,206.78** | **\$32,120.72** | **\$77.67** | **\$25.17** | **\$112.58** | **\$31.46** |

Note: SOC, standard of care; 2wT, two-way texting

As shown in Table 3, expanding 2wT to all new ART clients at MPC (scenario 1) with the assumption that 44% would enroll in 2wT, would decrease the total costs by 63.8% and 12.1% for SOC and 2wT, respectively. The increase in SOC cost was driven primarily by personnel costs, as more EC personnel would be needed for more clients. For 2wT, there was only a minor increase in the 2wT data officer FTE, increasing to 100%. Expanding to four other LH clinics (scenario 2), as shown in Table 5, would increase the cost per client by 277.2% for SOC, but decrease by 12.4% for 2wT. The main cost driver for SOC was personnel, utilities costs and fixed costs. The 2wT costs slightly decreased due to reduced personnel costs and no utilities costs. In scenario 2, with 2wT scaling up, 2wT the unit cost decreases.

### Incremental Cost-Effectiveness Ratio (ICER)

Table 6 describes the ICER in the scenario analyses. In the base case scenario with 468 participants in each group, SOC costs $33,458.72 with a unit cost of $107.58 and 2WT costs $36,670.38 with a unit cost of $98.05, resulting in a higher ICER for SOC. However, in a scenario where 2WT is extended to all new ART patients at MPC (N=2678), the unit cost drops significantly to $54.53 (total cost $54,807.37) for SOC, and $43.65 (total cost of $41,116.44) for 2WT. This shift results in a substantial decrease in ICER for 2WT, transitioning from +$24,705 to -$105,315(500% lower), indicating cost savings. Finally, in the second scenario where 2WT is implemented at four other LH clinics (N=2901), it becomes even more cost-effective with a unit and total cost of $31.46 and $32,120.72, respectively, and a lower ICER, resulting in substantial cost savings of -$723,739 compared to SOC.

**Table 6:**
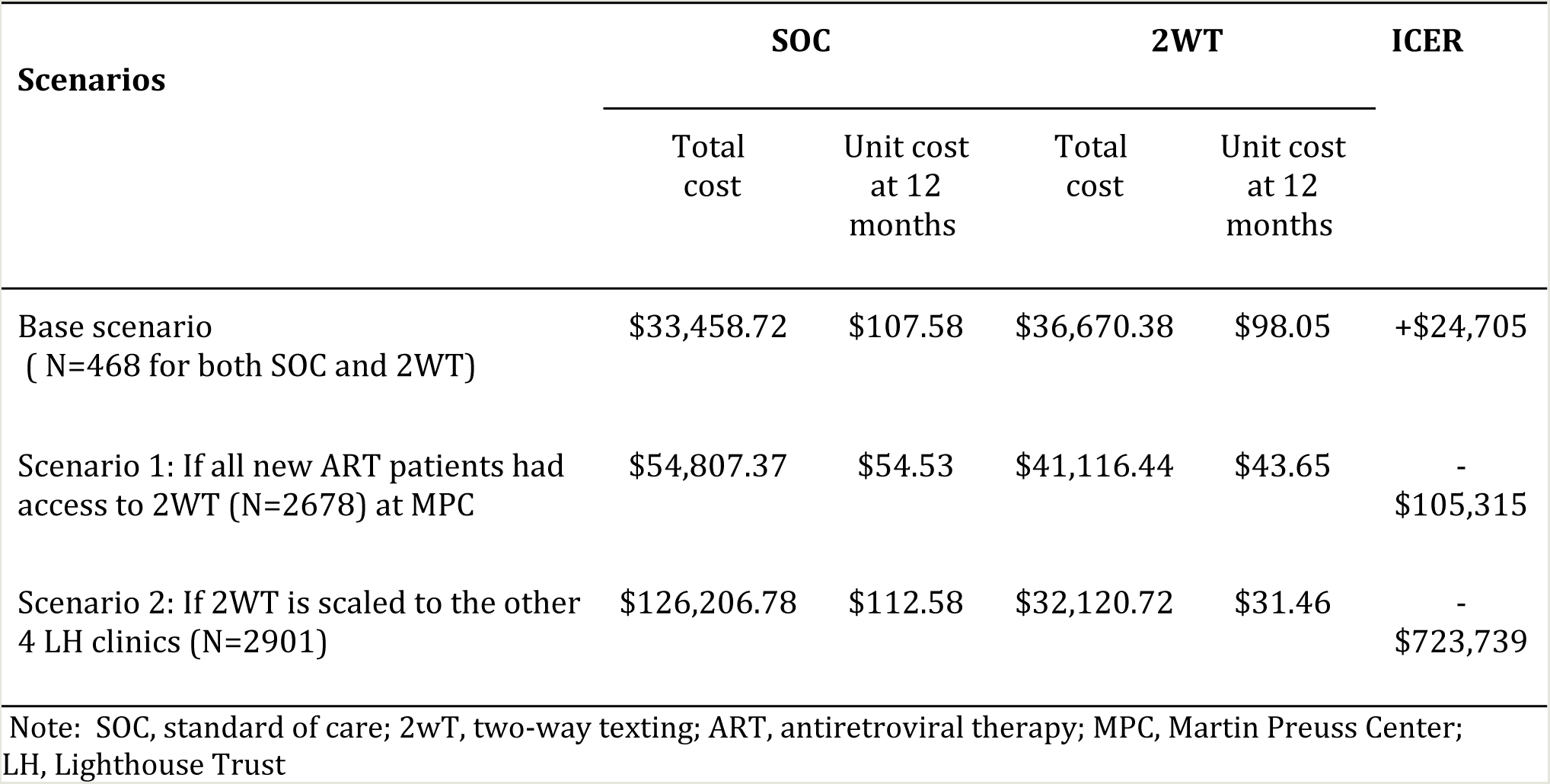
Change in cost and ICER with different scenarios.

## Discussion

We conducted a comprehensive cost-effectiveness analysis based on a payer costs (program/organization level) to evaluate the implementation of 2wT mHealth intervention for ART retention among newly initiated clients in Malawi, comparing it with the SOC. Our analysis underscores the cost-effectiveness of the 2wT intervention when implemented on a large scale. Our previous results indicated that the 2wT intervention yielded higher retention rates (80%) compared to SOC (67%) 12 months post-ART initiation. The total annual costs for 2wT were slightly higher than SOC costs. Personnel costs constituted the largest expense for 2wT, while utilities costs were the highest for SOC. Although the cost per client enrolled was higher for 2wT, the higher client retention 12 months post ART initiation resulted in a lower cost per client retained in care after 12 months compared to SOC.

We also explored two scenarios for our sensitivity analyses: one examining the expansion of 2wT to all new ART clients and another considering its implementation across LH facilities. Our study results indicated that, as 2wT expanded, its unit costs decreased. Furthermore, the 2wT intervention was associated with a positive ICER compared to SOC. Expanding 2wT for all new ART initiates at MPC significantly reduced the unit cos, further expansion to four more clinics reduced costs dramatically. In either scenario, the ICER had negative values, indicating improved outcomes and cost savings. These findings collectively emphasize the potential advantages of the 2wT intervention across diverse scenarios, showcasing improved patient retention and suggesting the feasibility of achieving cost savings through strategic implementation.

The findings of this cost-effectiveness analysis support the use of the 2wT mHealth intervention as a cost-effective method for improving ART retention in a large public HIV clinic in Malawi. The 2wT was deliberately built using an open-source CHT app, a global digital good, and as such, there are no licensing fees associated with the app use. The absence of licensing fees contributes to the reduction in the total cost of ownership, a key consideration for MOH decision-making process regarding potential national-scale implementation of the 2wT. Also in previous research, the 2wT intervention has been shown to be effective, usable and acceptable [34] among new ART initiates for improving early retention at MPC. The 2wT format confirms delivery of SMS sent, and it allows the client to respond at their convenience and discreetly, whereas unanswered phone calls may not register as missed calls to the recipient, and there are no voicemail options for Malawi phone plans. The 2wT is also free, thereby encouraging client feedback. Additionally, the 2wT intervention is also beneficial to the healthcare facility by helping HCWs streamline their workload by reducing unnecessary tracing and automating the generation of defaulter lists, which can lead to improved efficiency and resource allocation within the healthcare system [37].

The 2wT was also found to be beneficial in other contexts for other HIV interventions where HCWs found the system to be usable and encouraged clients to engage in their care [25,38]. However, potential challenges to scaling up were acknowledged, including the requirements for practice-based mentoring and constant supervision, generating client demand for 2wT, enrolling clients beyond ART initiation visits, establishing a supportive national policy enabling minors to enroll with guardian support, and addressing reporting redundancies to enhance efficiency.

The 2wT intervention was implemented to improve routine services [34]. Given the substantial cohort of nearly 26,000 ART-enrolled clients at MPC, coupled with an annual influx of approximately 2000 new ART clients, we recommend that 2wT be used to supplement the existing early retention program, emphasizing the importance of ensuring minimal additional workload for HCW for optimal effectiveness. In addition, robust training and mentorship for proper implementation is imperative. It is noteworthy that because HCWs at MPC were involved in the development of the 2wT system, they may have greater buy-in-a dynamic that may differ when scaling the system to other LH facilities when the system is already developed. Implementing mHealth interventions for ART adherence at scale is feasible and cost-effective. These interventions can build on existing healthcare infrastructure and leverage available resources [5].

Our study provides crucial insights into the cost implications of implementing a mHealth intervention within a routine setting. Notably, the early retention program at MPC is just one piece of a larger effort to keep patients engaged in care. MPC offers a wide range of HIV care and treatment services across the HIV cascade [29,30]. Because of this setup, at MPC, the HCWs have varied roles and responsibilities that often cut across different departments. As HCWs take on more senior positions, their roles become even more complex and overlapping, adding challenge to accurately calculate the specific costs associated with the early retention services within the regular program. Thus, in addition to the time-motion study, we had to continuously refine our estimates with the HCWs involved in retention services to accurately estimate the different cost categories. MPC has detailed records for documenting clinic visits through the EMR and individual patient files.

### Limitations

It is essential to acknowledge some limitations of this study. Firstly, not all clinic operational costs, such as senior leadership personnel expenses, were incorporated, potentially leading to an underestimation of total costs. The SOC costs were extracted from the overall MPC program expenditure data; the absence of specific SOC intervention-based expenditure recording within LH accounting records could have omitted certain costs. Utility or building costs could vary more than estimated. Furthermore, in 2020, EC records were incomplete. Consequently, the frequency of EC interactions with the clients and referrals to back-to-care could not be completely ascertained. Lastly, the 2wT is limited to those who have exclusive use of a mobile phone and are literate. This highlights the need to consider 2wT as an augmentation or complement to SOC and not a replacement. 2wT is not yet integrated into the underlying MPC electronic medical record system; however, planned integration should further lower costs through more automated visit reminders and referrals to B2C. Despite these limitations, the 2wT intervention should still be considered an effective intervention for those who can use it, as it has demonstrated positive outcomes in improving ART retention.

## Conclusion

Based on this study, we conclude that 2wT is a cost-effective method to improve early retention of new ART initiates in Lilongwe, Malawi. 2wT reduced cost per retained client when compared to those receiving the SOC. Our scenario analyses demonstrate that as the 2wT is expanded in scale, it may yield cost savings, thereby establishing its cost-effectiveness for the clinic. The 2wT intervention should not replace the existing SOC services, rather, it should complement the existing retention interventions to support engagement in care. This study sets the stage for future investigations aimed at assessing the cost-effectiveness of scaling up the 2wT program, measuring 2wT impact on timely visit attendance, client retention, attrition, and reengagement in care over time. The results of this study will also be shared with the MOH, and other key stakeholders to advise on how 2wT may be expanded to other facilities, including government facilities, as the MOH considers using mHealth interventions, like 2wT, to strengthen the national HIV program [12].

## Data Availability

All data produced in the present study are available upon reasonable request to the authors.

## Acknowledgments

Research reported in this publication was supported by the Fogarty International Center of the National Institutes of Health (NIH) under Award Number R21TW010583 (CF), and PATH under Award Number AID-OAA-A-16-00084. The content is solely the responsibility of the authors and does not necessarily represent the official views of the National Institutes of Health or PATH. The authors would also like to thank the Lighthouse Trust retention program and research department for their partnership in implementing the 2wT intervention.

## Authors’ Contributions

CKK: Writing – original draft. CKK and HW: Formal Analysis. CKK, JS, HC, AT, OS, GC, JC and JH: Investigation and Project Administration. CKK, MC, HW, and HC: Data curation. CF, WG, MP and HT: Supervision.HT and CF: Conceptualization. All authors: Writing – review & editing.

## Appendix

## Appendix 1: Data Collection Tools for time-motion study

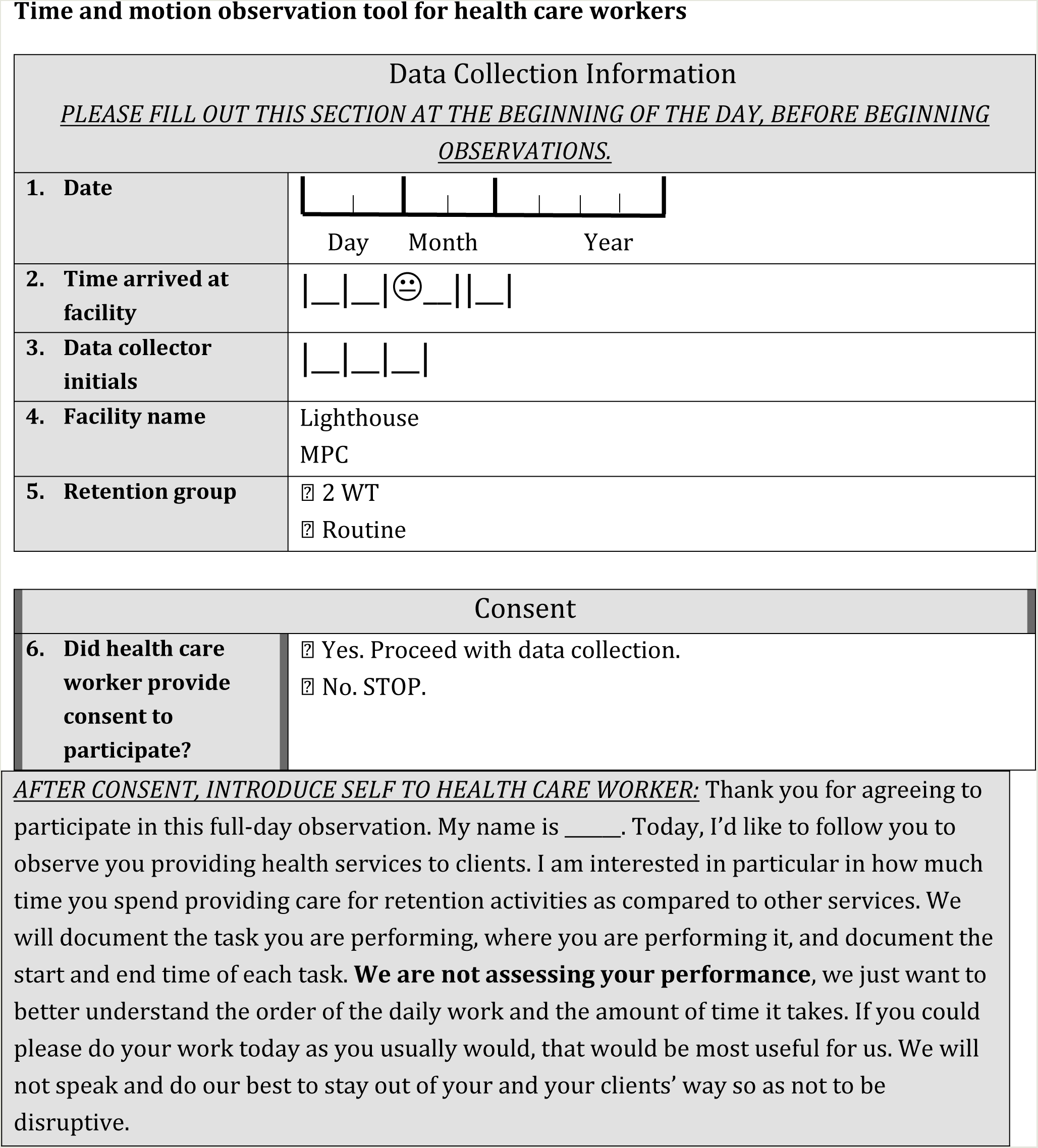
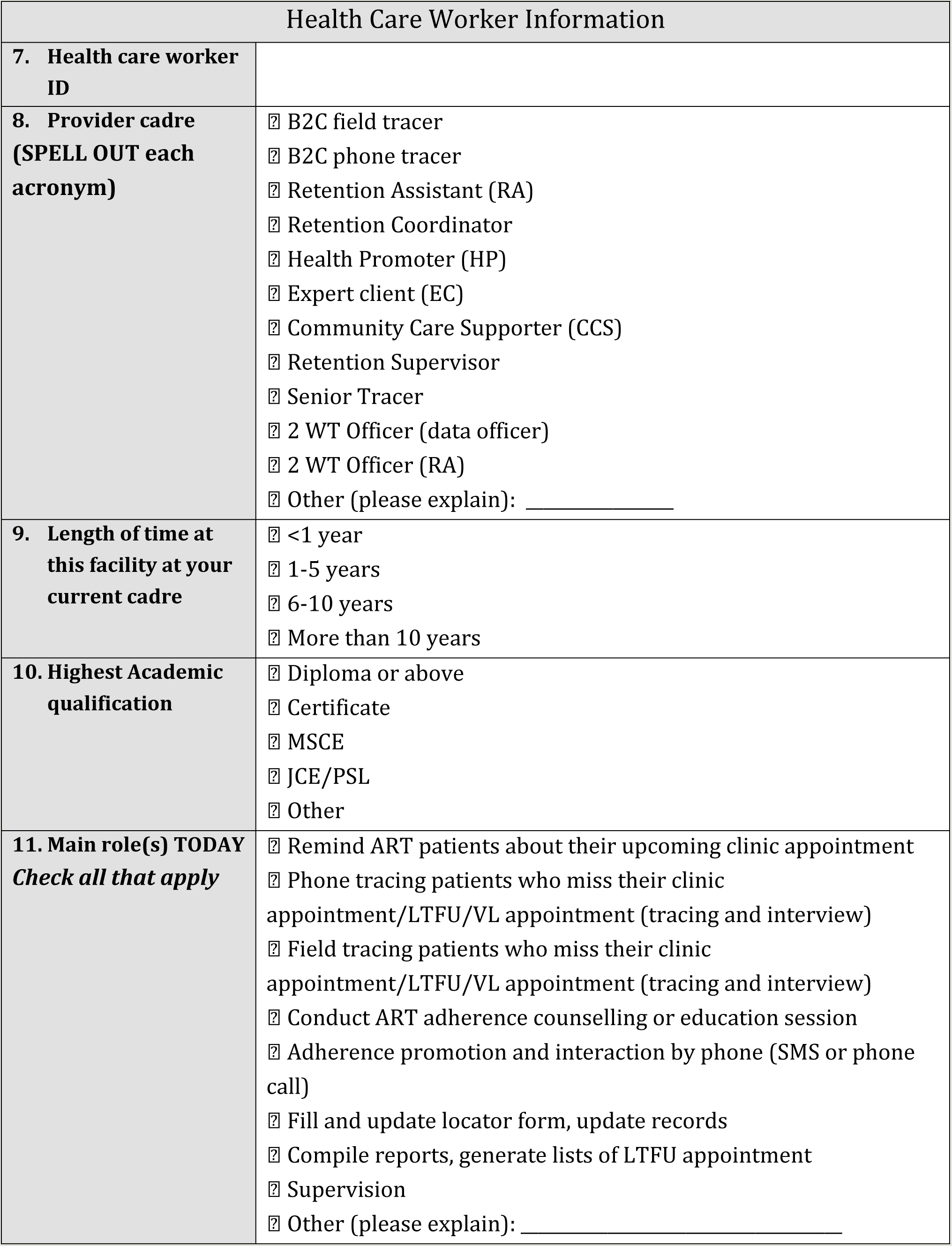

## Appendix 2: Consolidated Health Economic Evaluation Reporting Standards 2022 (CHEERS 2022)

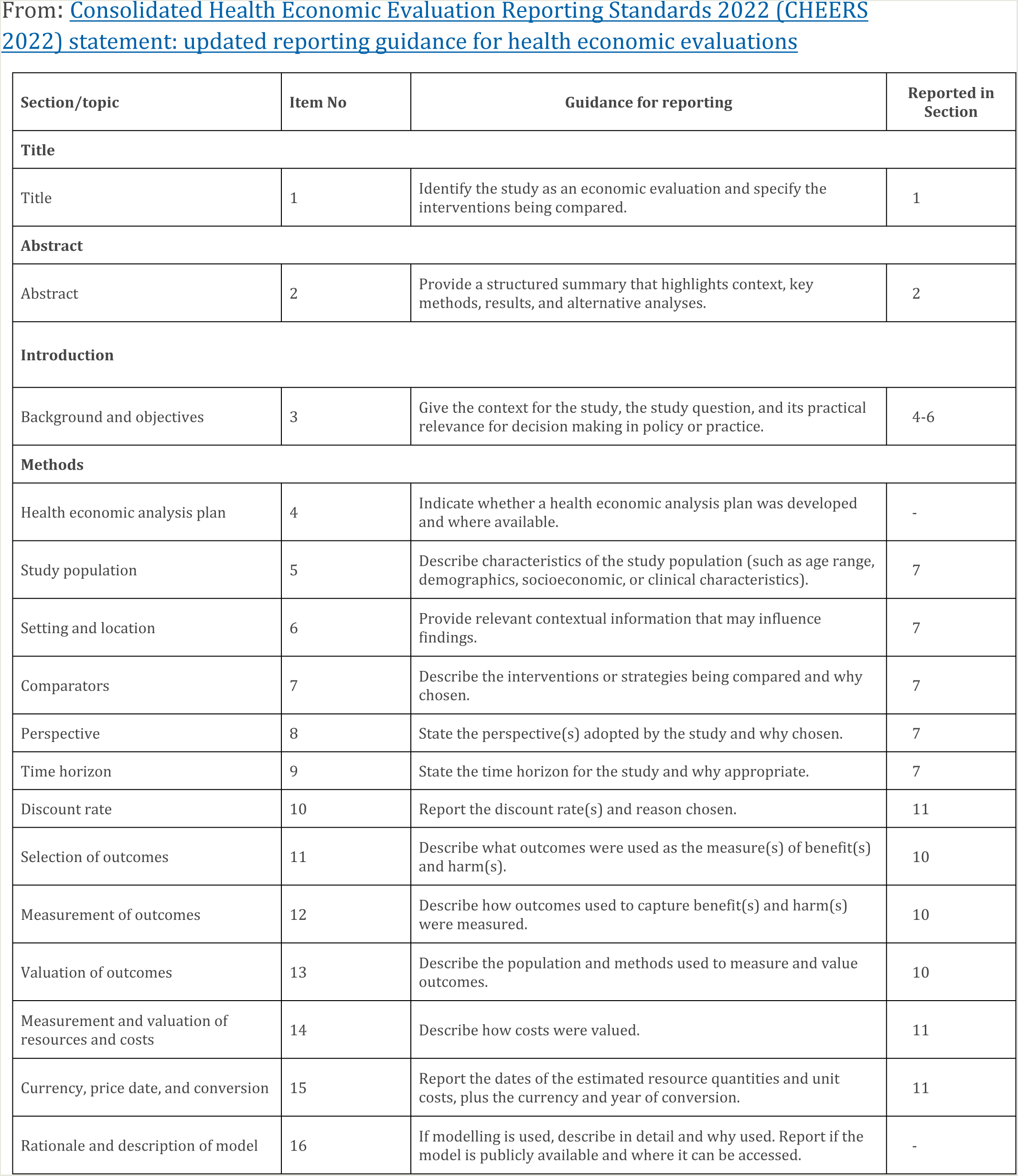
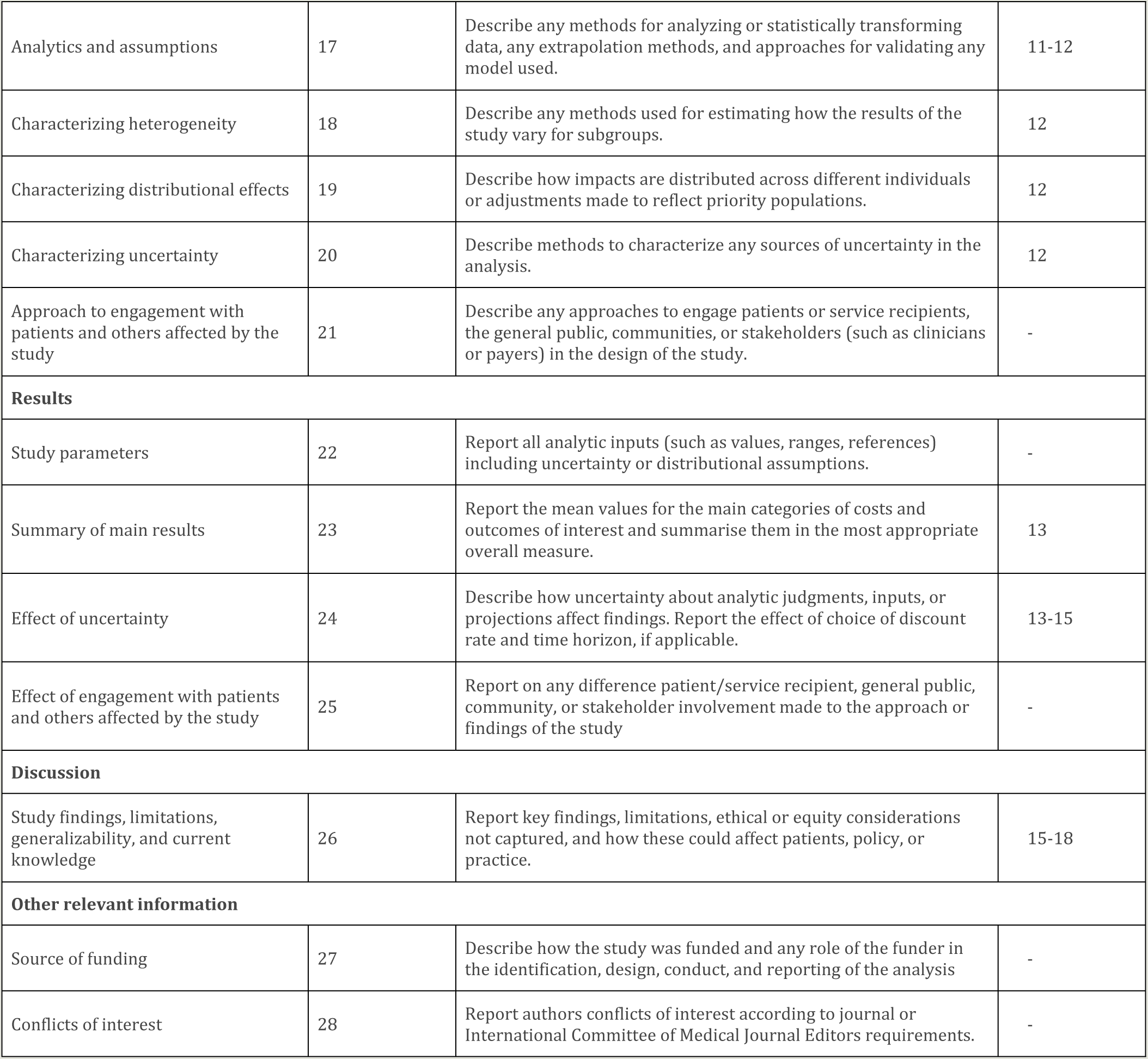

## Appendix 3: Cost shares by input categories for all costs

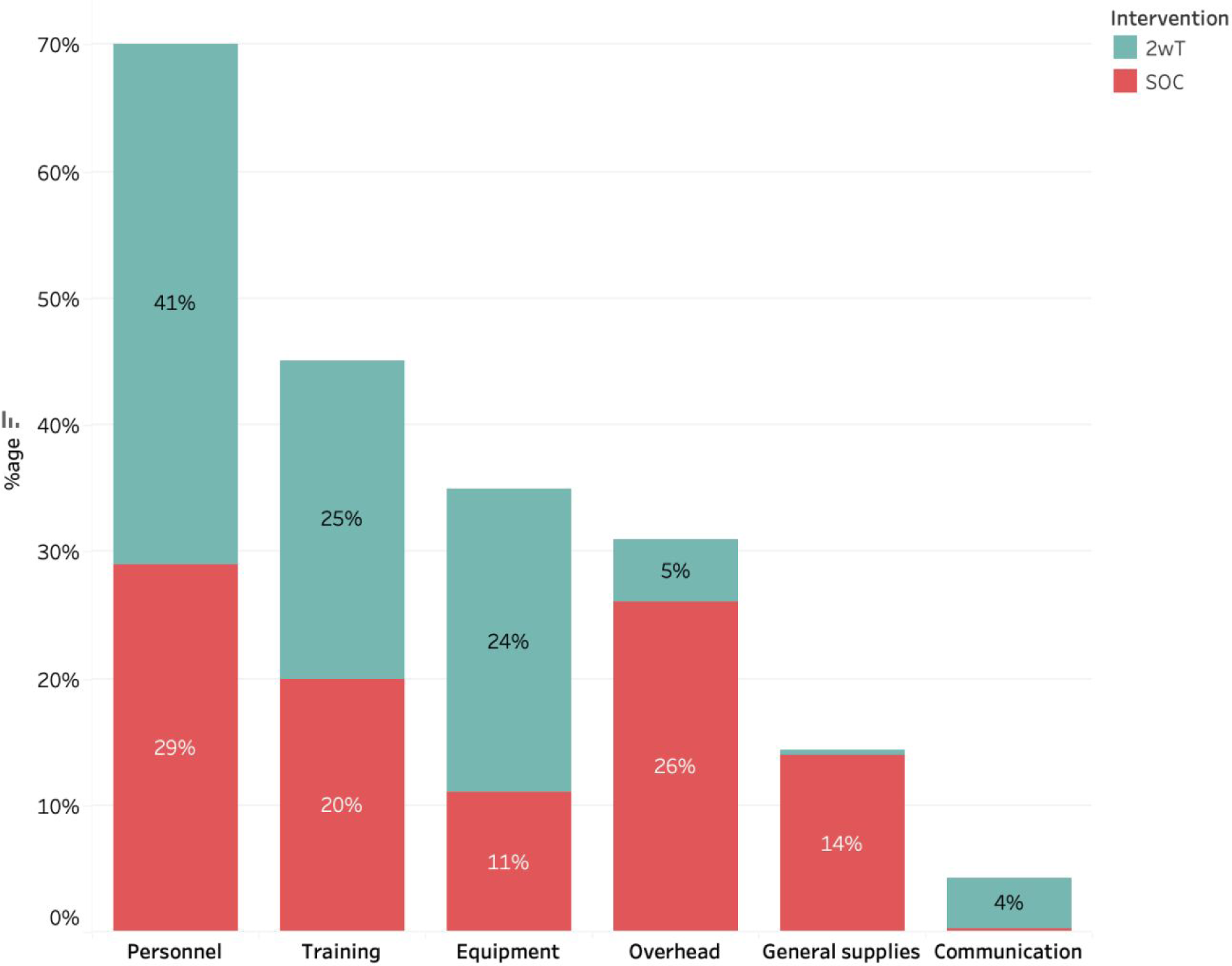

